# Optimized Flow Cytometric Detection of Transient Receptor Potential Vanilloid-1 (TRPV1) in Human Hematological Malignancies

**DOI:** 10.1101/2021.08.04.21261521

**Authors:** Sofia A Omari, Dominic P Geraghty, Alhossain A Khalafallah, Pooja Venkat, Yvette M Shegog, Scott J Ragg, Charles E. de Bock, Murray J Adams

## Abstract

The ectopic overexpression of transient receptor potential vanilloid-1 (TRPV1) has been detected in numerous solid cancers including breast, prostate, pancreatic, and tongue epithelium cancer. However, the expression of TRPV1 in hematological malignancies remains unknown. Here we show through *in silico* analysis that elevated TRPV1 mRNA expression occurs in a range of hematological malignancies and present an optimized flow cytometry method to rapidly assess TRPV1 protein expression for both cell lines and primary patient samples. Three anti-TRPV1 antibodies were evaluated for intracellular TRPV1 detection using flow cytometry resulting in an optimized protocol for the evaluation of TRPV1 in hematological malignant cell lines and patients’ peripheral blood mononuclear cells (PBMC). Overexpression of TRPV1 was observed in THP-1 (acute monocytic leukemia) and U266B1 (multiple myeloma, MM), but not U937 (histiocytic lymphoma) compared to healthy PBMC. TRPV1 was also detected in all 49 patients (including B-cell non-Hodgkin’s lymphoma (B-NHL), MM and others, and 20 healthy controls. TRPV1 expression was increased in 8% of patients (MM=2, B-NHL=2). In conclusion, we provide an optimized flow cytometry method for routine expression analysis of clinical samples and show that TRPV1 is increased in 8% of patients with hematological malignancies.

## Introduction

The transient receptor potential vanilloid-1 (TRPV1) protein is a transmembrane, non-selective cation channel activated by a variety of stimuli, including heat, acidic pH, capsaicinoids and other plant-derived vanilloids (Caterina and Julius 2001). Although renowned as a nociceptive receptor (Holzer 2008), TRPV1 expression has been detected in many cancers including glioma (Contassot et al. 2004a), breast cancer (Weber et al. 2016), prostate cancer (Sánchez et al. 2005), colon adenocarcinoma (Domotor et al. 2005) and pancreatic cancer (Mergler et al. 2012). Significantly, capsaicin, a vanilloid and TRPV1 agonist, was found to be cytotoxic to a number of cancer cell lines through both TRPV1-dependent and -independent mechanisms raising the prospect that TRPV1 expression can be exploited therapeutically (Bao et al. 2019; Basith et al. 2016; Fattori et al. 2016). To date, there has only been a limited assessment of TRPV1 expression in either normal blood cells or hematological malignancies (Omari et al. 2017). TRPV1 has been detected in rat blood neutrophils (Wang et al. 2005), microglia, the brain-resident macrophage (Kim et al. 2006), and human blood leukocytes (Spinsanti et al. 2008; Saunders et al. 2007). TRPV1 protein was also detected in human dendritic cells (DC) as it plays a role in DC maturation (Toth et al. 2009), while there remains controversy regarding TRPV1 protein and mRNA expression in mice DC (O’Connell et al. 2005; Basu and Srivastava 2005). The activation of TRPV1 and its role in THP-1 monocyte migration and adhesion, and U937 cell apoptosis have been previously studied (Maccarrone et al. 2000; Himi et al. 2012; Schilling and Eder 2009). Recently, Punzo et al reported that the vanilloid resiniferatoxin, an ultrapotent TRPV1 agonist, produced pro-apoptotic and anti-proliferative signals in primary lymphoblasts from acute T-lymphoblastic leukemia patients. They also detected TRPV1 mRNA in Jurkat cells and patient lymphoblasts (Punzo et al. 2018). Moreover, treatment of the hematological cells lines THP-1, U266B1 and U937 *in vitro* with capsaicin causes apoptosis (Omari et al. 2016). Given that TRPV1 is a transmembrane protein and the availability of commercial anti-TRPV1 antibodies, flow cytometry is the ideal method for detection. However, previous studies using flow cytometry to analyze TRPV1 expression suffer from insufficient negative or isotype controls, or appear to omit permeabilization steps when using antibodies that target intracellular antigens (Caprodossi et al. 2011a; Huang et al. 2010; Amantini et al. 2004; Amantini et al. 2007; Amantini et al. 2009). Similarly, non-specific binding was reported with immunohistochemistry using the same antibodies commonly used for flow cytometry when testing TRPV1^-/-^ mice bladder tissue, resulting in false positive results (Everaerts et al. 2009). This demonstrates that the expression of TRPV1 needs to be re-evaluated.

## Materials and Methods

### *In silico* RNA-seq data analysis

To determine *TRPV1* gene expression levels, *TRPV1* in total RNA obtained using next-generation sequencing was extracted from published RNA-seq datasets collected originally for clinical trials including BEAT AML (n=224), TCGA-LAML (n=151), TARGET-AML (n=145), TARGET-ALL-P2 (n=441), TARGET-ALL-P3 (n=78), TCGA-DLBC (n=48), CTSP-DLBCL1 (n=41), and CGCI-BLGSP (n=109). *TRPV1* gene expression was plotted as fragments per kilobase of transcript per million mapped reads (FPKM) for clinical data, and transcript per million (TPM) count using GraphPad Prism© (v 8.4.2, San Diego, CA, USA). Tukey’s multiple comparisons test (One-way ANOVA) was used for statistical analysis.

### Patient Blood Collection

Peripheral whole blood was collected from healthy adult donors and patients with hematological malignancies using sterile venipuncture. Blood was collected into EDTA (for flow cytometry), and CPT Vacutainer^®^ tubes (BD Biosciences, San Jose, USA), to isolate patients PBMCs for Western blot. The study was approved by the Human Research Ethics Committee Network, Tasmania (H0011050).

### Cell Culture

THP-1 cells (ECACC, UK) were cultured in RPMI1640 (Life Technologies, Grand Island, USA) supplemented with 20% heat-inactivated fetal bovine serum (Life Technologies), 2 mM glutamine, 100 U/mL penicillin and 100 µg/mL streptomycin. U266B1 and U937 cell lines (ATCC, Manassas, VA, USA) were cultured in RPMI1640 containing 2 mM L-glutamine, 10 mM HEPES, 1 mM sodium pyruvate, 4.5 g/L glucose, 1.5 g/L sodium bicarbonate and supplemented with 15% and 10% FBS, respectively.

### Flow cytometry protocol development

Three polyclonal rabbit anti-TRPV1 antibodies [SC-20813, purified by proprietary techniques (Santa Cruz Biotechnology, CA, USA); ACC-030, affinity purified on immobilized antigen (Alomone Labs, Jerusalem, Israel); LS-C150735, affinity purified (Lifespan Biosciences, WA, USA), and polyclonal IgG goat anti-rabbit secondary antibody-FITC (Santa Cruz Biotechnology) were initially assessed. Testing of fixation and permeabilization kits [CALTAG™ (Life Technologies); Cytofix/Cytoperm™ (BD Biosciences)], blocking agents [AB serum (from a healthy donor), 10% goat serum (Life Technologies), FcR blocking reagent (Miltenyi Biotechnology, Cologne, Germany) and a mixture of 10% human AB serum, 1% BSA and 0.05% sodium azide, and goat serum mix (10% goat serum + 0.1% BSA + 0.05% sodium azide)] was also performed. Isotype control rabbit IgG, sc-3888 (Santa Cruz Biotechnology) was used as a negative control in a same quantity/dilution as the primary anti-TRPV1 in all experiments. Exclusion of TRPV1 antibody was used as an additional negative control. Protocols were developed to produce the best signal (specific binding) to noise (non-specific binding) ratio for Western blotting and separation of TRPV1 signal from isotype control for flow cytometry.

### Western Blotting

Cultured cells or patients PBMCs were lysed, protein extracted, and the concentration measured as previously described (Saunders et al. 2007). Protein samples were diluted 1:1 with Laemmli buffer-2-ME. Samples were then heated for 5 min at 95°C and loaded onto SDS-PAGE gels (Mini-PROTEAN^®^ TGX™ Precast Gels, Bio-Rad Laboratories, CA, USA). PageRuler™ plus prestained protein (Fermentas, Thermo Scientific, Burlington, Canada) and MagicMark™ XP (Life Technologies) were used as ladders. The protein samples were electrophoresed at 200 V for 45 min using a running buffer containing 25 mM Tris, 192 mM glycine, 0.1% SDS, pH 8.3 (Bio-Rad Laboratories, CA, USA) at 4°C. Protein bands were transferred to PVDF membranes at 95 V for 1 h at 4°C. The membranes were blocked with 3% BSA, 5% non-fat milk, 0.05% Tween-20 in phosphate buffered saline (PBST), for 1 hr at RT on an orbital shaker. The membranes were then washed for 5 min with PBST and incubated at 4°C overnight with rabbit polyclonal anti-TRPV1 (LifeSpan Biosciences, WA, USA) at 1:10000 in the blocking buffer. The blots were then washed with PBST for 5 min (4X) and incubated for 1 h with goat anti-rabbit IgG-HRP (Cell Signalling Technology, MA, USA) at 1:5000 in the blocking buffer then washed. The membranes were developed with chemiluminescence using the Immobilon™ Western detection kit (Millipore, MA, USA) for 5 min and visualized using LAS-3000 Image reader (Fuji, Tokyo, Japan). Membranes were re-washed, re-blocked for 20 min, and re-incubated for 2 hr with rabbit monoclonal anti-human GAPDH (Cell Signaling Technology) at 1:3000 in the blocking buffer. Membranes were subsequently washed, stained and visualized. Forty micrograms of protein lysate were loaded for all control and patient samples. THP-1 cells were used as a positive control (Himi et al. 2012).

### Flow cytometry

One million cells were surface stained with CD45-PerCP (clone 2D1), and CD3-Brilliant Violet™ 421 (clone UCHT1), or CD14-V500 (clone M5E2), and CD19-PE (clone HIB19), all from BD Biosciences, and incubated for 10 min in the dark at RT. Erythrocytes were then lysed for 10 min at RT using FACS Lyse solution (BD Biosciences). Cells were centrifuged at 350 x g for 5 min, then washed twice in PBSA (10% FBS, 0.1% sodium azide in PBS). Cells were fixed for 20 min at 4°C using BD Cytofix/ Cytoperm™ kit, then washed twice using the permeabilization buffer. Non-specific binding sites were blocked for 15 min at RT using 10% AB human serum, 1% BSA, 0.05% sodium azide in the permeabilization buffer. Cells were then intracellularly stained with 0.5 µg of rabbit polyclonal anti-TRPV1 (Lifespan Biosciences) or 0.5 µg (1:25) of isotype control for 45 min at 4°C. After washing, cells were stained with secondary antibody (1:25) for 20 min at 4°C and washed twice. Finally, cells were resuspended in Ca^2+^/ Mg^2+^ free PBS prior to acquisition on the Attune^®^ Acoustic Focusing Cytometer (Applied Biosystems^®^/Life Technologies, Grand Island, USA). FlowJo™ software (OR, BD, USA) was used to analyze the data. A flow chart of the protocol can be found in Supplementary Fig. S1.

Samples from 49 patients and 20 age- and sex-matched healthy subject controls with no history of malignancies, were collected and characterized (Table S1). Control samples were run simultaneously and analyzed with patient samples for each experiment. Patients diagnosed with myeloproliferative disorders (MPD) included chronic myelocytic leukemia (CML), chronic myelomonocytic leukemia (CMML) and essential thrombocythemia (ET) (n=1 each). Median fluorescent intensity (MFI) ratio was generated by dividing MFI (patient cells) by MFI (control). The TRPV1 expression was then designated as increased (MFI ratio ≥2), decreased (MFI ratio <0.5) or similar (MFI ratio 0.5-1.9).

## Results

### TRPV1 is expressed in subsets of hematological malignancies including AML, ALL and DL-BCL

To determine the expression of *TRPV1* in hematological malignancies, an *in silico* analysis was carried out across a range of clinical RNA-seq data sets and revealed significantly higher expression of *TRPV1* in AML and ALL patients within TCGA-LAML, TARGET-ALL-P2 and TARGET-ALL-P3 compared to BEAT AML, TARGET_AML, TCGA-DLBC, CTSP-DLBCL1 and CGCI-BLGSP (P <0.0001) (Fig. 1A).

**Fig. 1.**
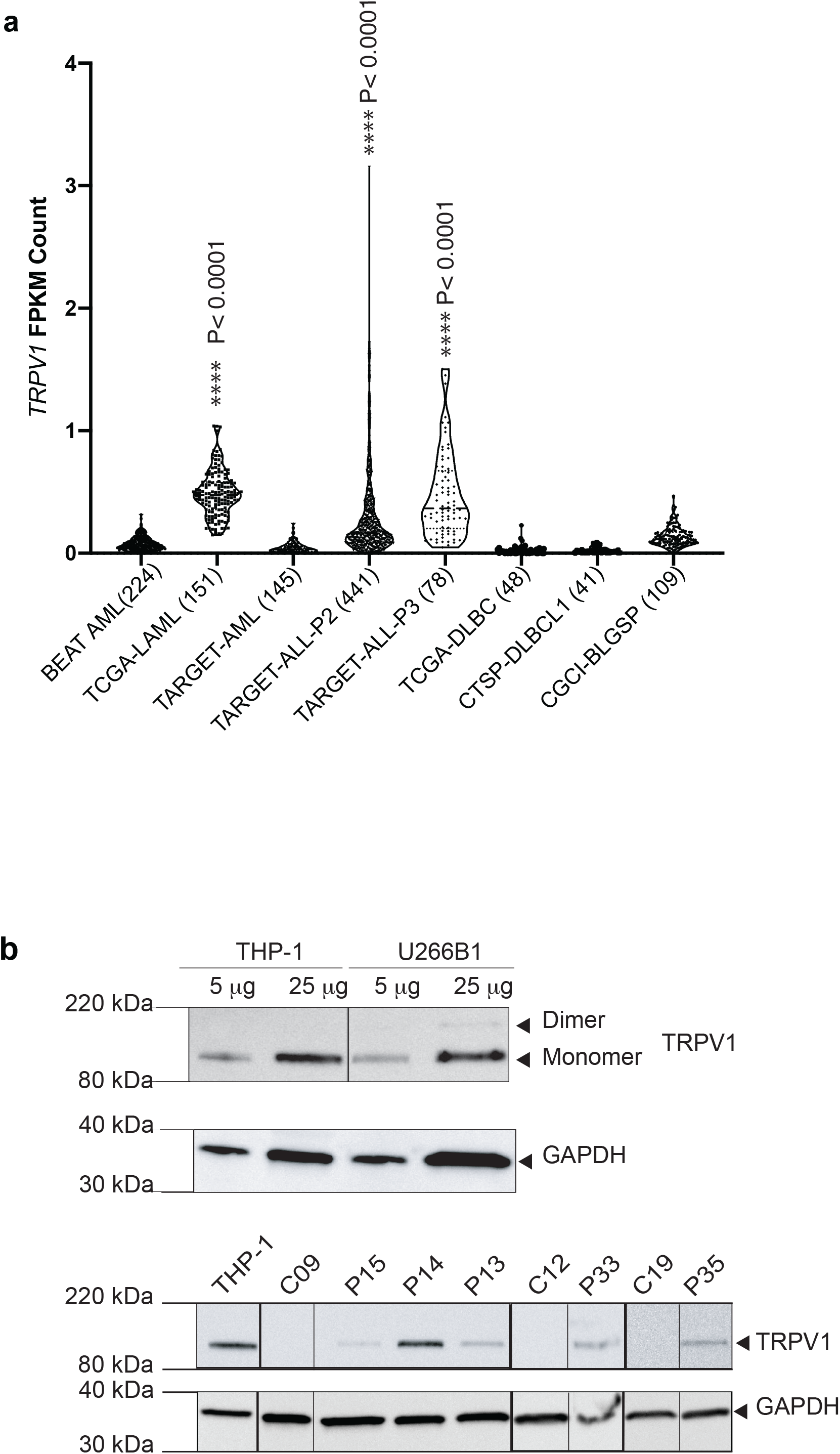
*TRPV1* gene and protein expression using RNA-sequencing data and Western blotting. (A) *TRPV1* gene expression in different databases. *P<0.0001 compared to all others (Tukey’s). (B) TRPV1 expression detected using Western blotting in THP-1 and U266B1 cell lines and PBMCs protein samples of patients with hematological malignancies. THP-1 protein was used as a positive control, and GAPDH was used as an internal control. TRPV1 monomer (∼ 95 kDa) was detected in five patients (P#13, P14, P15, P33 P35)

We next sought to validate this expression at the protein level. Western blotting was first performed on THP-1 cells and U266B1 myeloma cells that express TRPV1 monomer (∼95kDa) and U266B1 that also expresses a dimer (Fig. 1B). This was then extended to a range of different patient samples, with TRPV1 detected in some patients (MM=1, B-NHL=4) (Fig. 1B). However, for routine clinical analysis of protein expression, flow cytometry is preferred.

Optimized intracellular flow cytometry analysis can robustly determine TRPV1 expression in clinical samples. The majority of commercially available TRPV1 antibodies target intracellular epitopes. Some studies that employed a flow cytometric method to determine TRPV1 expression did not describe permeabilization step or include appropriate isotype control which affect the validity of the result (Basu and Srivastava 2005; Caprodossi et al. 2011b; Amantini et al. 2007). To this end, three commercial anti-TRPV1 antibodies were assessed that bind different intracellular regions at the C- and N-termini (Fig. 2A). Both the rabbit anti-TRPV1 antibody (Santa Cruz Biotechnology, SC-20813) and the rabbit anti-human TRPV1 antibody (Alomone Labs, ACC-030) were not able to distinguish TRPV1 expression over isotype controls (Fig. 2B,C), even when alternative blocking methods, including 10% goat serum, human AB serum or FcR blocking reagents were used (data not shown). The rabbit anti-TRPV1 antibody (Lifespan Biosciences, LS-C150735) reliably detected TRPV1 expression with excellent separation of the TRPV1 signal from the isotype control and revealed a quantitative differential expression in cell lines: THP-1> U266B1>U937, with the latter being comparable to healthy control PBMCs (Fig. 2D, and Table S2). Median fluorescence intensity (MFI) within healthy PBMC subpopulations, was: CD19-B-cells (1.36×10^5^) > CD14-monocytes (1.21×10^5^) > CD3-T-cells (0.96×10^5^). Using this optimized protocol and staining methods, we analyzed 49 primary patient samples^4^.

**Fig. 2.**
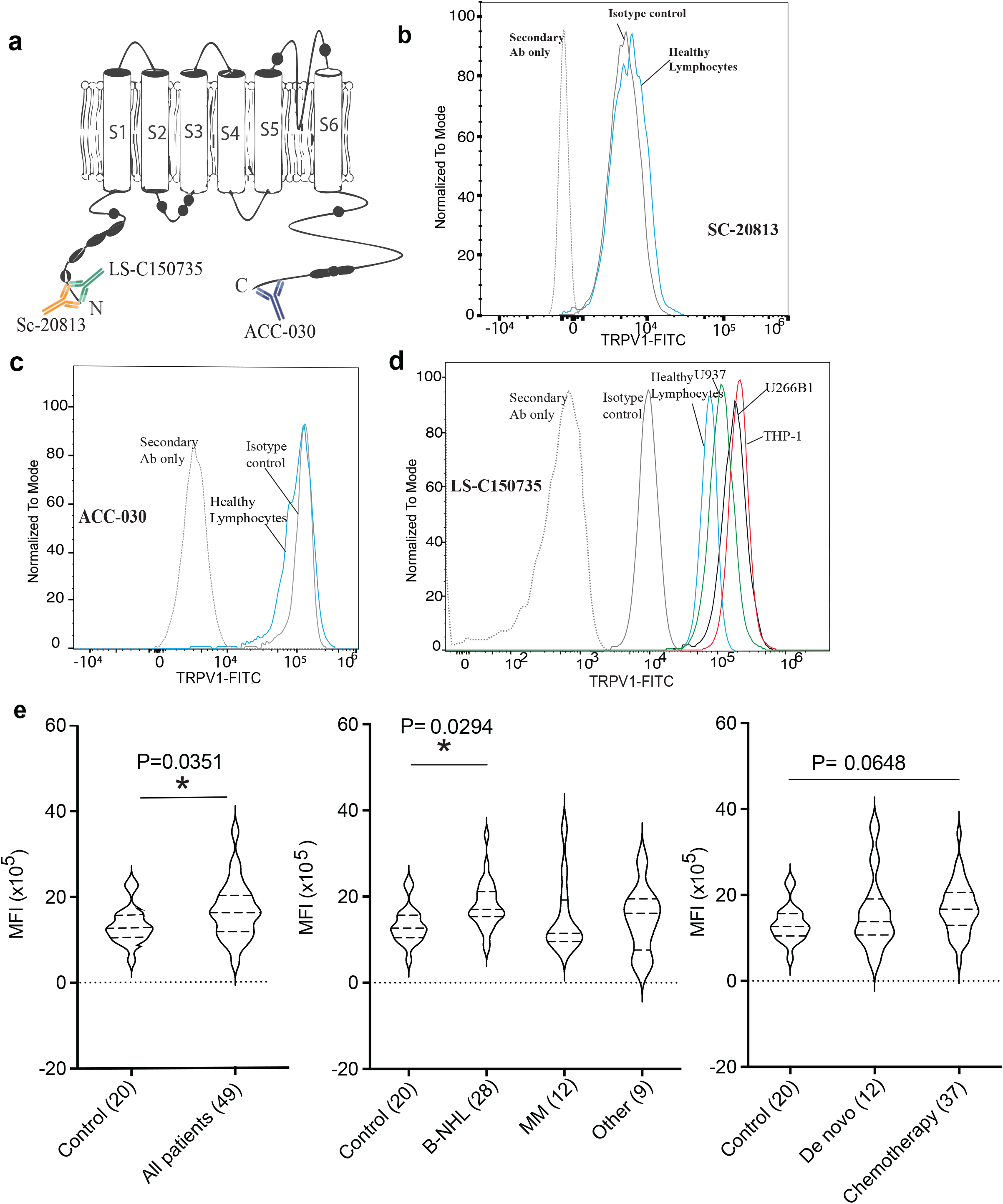
TRPV1 expression using Flow Cytometry. (A) Overview structure of TRPV1 and the 3 antibodies binding sites. Assessment of anti-TRPV1 antibodies: (B) SC-20813, (C) ACC-030, and (D) LS-C150735. Panel D demonstrates clear separation of isotype control from TRPV1 signals in THP-1, U266B1, U937 cells and healthy human lymphocytes using LS-C150735. (E) TRPV1 expression in all patients with hematological malignancies and healthy controls (n=20). TRPV1 in MM, B-NHL and other hematological malignancies group (treated and *de novo*) were compared to control. Median fluorescence intensity (MFI) value for individual patients expressed using violin plots with the mean and standard deviation also shown. Patients samples were also expressed as a ratio (MFI patient: MFI control). Overall TRPV1 was significantly higher in all patients with hematological malignancies compared to healthy control (P=0.0351, unpaired t-test) with B-NHL patients having significantly higher TRPV1 compared to control (P= 0.0294, Dunnet’s test)

Based on the distribution of the MFI ratio for all patients, ectopically high TRPV1 expression was deemed to be an MFI of 2-fold over normal healthy controls. Using this cut off, increased TRPV1 expression was detected in 8.2% of patients (4/49). Of these, two had MM and two B-NHL. Of the 24 under-treatment and four *de novo* B-NHL patients, TRPV1 was found to be similar to healthy control in 26 patients. However, TRPV1 was increased in 2 females diagnosed with diffuse large B cell lymphoma, who were under treatment (one with radiation only, and the latter on Rituximab). Furthermore, with the nine under treatment and three *de novo* MM patients, TRPV1 expression was found to be similar (n=10), or increased (n=2, a *de novo* with lambda, B-2-microglobulinimia, and an under-treatment patient with Waldenstrom macroglobulinemia) compared to controls. TRPV1 in patients with other hematological malignancies (n=9) were all found to be similar to controls (Fig. 2E). The four patients (8.2%) with increased TRPV1-MFI ratios had the highest MFI values (mean± SD; 308783± 145474) relative to other patients (154178± 100242). C-reactive protein (CRP) values were elevated (≥ 5 mg/L) in 25% of the 44/49 patients measured, ranged from 1-106 mg/L.

## Discussion

This is the first study to report an optimized method to detect TRPV1 expression using flow cytometry in hematological malignancies. Numerous previous studies have analyzed TRPV1 expression during carcinogenesis (Gkika and Prevarskaya 2009). Overexpression of TRPV1 has been reported in solid cancers including glioma (Contassot et al. 2004b) and human tongue epithelium cancer (Marincsák et al. 2009), whereas lower expression was reported in urothelial cancer (Kalogris et al. 2010; Lazzeri et al. 2005). Capsaicin induced TRPV1-dependent apoptosis in some cancers, such as prostate (Ziglioli et al. 2009). Therefore, increased TRPV1 expression in blood cancers could create a new therapeutic target to treat these conditions. In the present study, a TRPV1 monomer (∼95 kDa), which represents the non-glycosylated form of the molecule (Vetter et al. 2006), was found to be strongly expressed in both THP-1 and U266B1 cells with a TRPV1 dimer (∼200 kDa) also detected in the latter and both with higher expression compared to healthy controls.

Significantly, we have for the first time established a robust flow cytometry protocol for the rapid evaluation of TRPV1 expression. The affinity purified anti-TRPV1 antibody (Lifespan Biosciences, LS-C150735) provided excellent separation of TRPV1 and isotype signals. This was in contrast to the affinity purified anti-TRPV1 antibodies from Alomone Labs and Santa Cruz Biotechnology which were unable to separate the TRPV1 signal from the isotype control, suggesting a lack of specificity for TRPV1. As TRPV1 was suggested to be ubiquitously distributed in all human cells (Cortright and Szallasi 2004; Fernandes et al. 2012), negative controls are warranted to distinguish a true expression of TRPV1 from false positive expression, with the ultimate negative control to test the specificity of the primary antibody would be the implementation of knockdown/knockout cells (Couchman 2009; Shim et al. 2007).

Using our optimized protocol, TRPV1 was detected in the PBMC fraction of all 49 patients with hematological malignancies, including B-NHL, MM and a range of other malignancies. TRPV1 expression was increased in ∼ 8 % of patients, but was not significantly different between MM and other hematological malignancy cohort, nor between *de novo* (untreated) and patients undergoing treatment. Differential expression of TRPV1 in subsets of patients with different hematological malignancies were consistent with the *in-silico TRPV1* gene expression data obtained from various clinical databases, with higher *TRPV1* expression in some of these groups compared to normal blood. One of the limitations of the present study is that TRPV1 expression was determined in single samples from individual patients who were at different stages of the disease and chemotherapy. Further studies of TRPV1 expression in a larger patient cohort and changes in TRPV1 expression over time, following chemotherapeutic intervention would allow what contribution altered TRPV1 expression, if any, plays in the pathogenesis, progression and chemotherapy-induced remission of individual hematological malignancies.

In conclusion, we present a robust, optimized flow cytometric method to measure TRPV1 expression and demonstrate increased expression of TRPV1 in THP-1 and U266B1 malignant hematological cells as well as in a subset of MM and B-NHL patients. The results from this study warrant further research into whether this may be exploited therapeutically using capsaicin or other vanilloids.

## Supporting information

Sup 1-TRPV1 FC protocol

Sup 3-Fig1B-raw data Western blot

## Data Availability

All data requests will be considered.

## Acknowledgements

The authors thank the Clifford Craig Foundation for generous financial support (G0018836), and the Australian Government for an Australian Postgraduate Award to SO.

## Compliance with Ethical Standards

### Research involving Human Participants

The study was approved by the Human Research Ethics Committee Network, Tasmania, Australia (H0011050).

### Informed consent

Informed consent was obtained from all individual participants included in the study.

## Declarations

### Availability of data and material

The authors confirm that the data supporting the findings of this study are available within the article and its supplementary materials.

### Code availability

Not applicable.

### Consent to participate

Informed consent was obtained from all individual participants included in the study.

### Consent for publication

Not applicable.

## Footnotes

4 *Supplementary data to be shared upon request*.

