## Supplementary material for "Optimized Flow Cytometric Detection of Transient Receptor Potential Vanilloid-1 (TRPV1) in Human Hematological Malignancies": Sup 1-TRPV1 FC protocol

Supplementary Fig. 1: overview of the step by step Flow cytometry protocol for the detection of TRPV1 in human leukocytes in peripheral blood

EDTA-peripheral blood (1 million WBCs)

↓

Surface staining (CD markers), incubate 10min, dark at RT

↓

RBCS lyses (5 min), then wash 2X with FACS buffer

↓

Fix cells, incubate 20 min, dark at 4C, then wash 2X with perm buffer

↓

blocking with 10% human AB type serum+1% BSA+ 0.05% NaN3 in perm buffer

↓ Incubate for 15 min, RT, then wash

Stain with 0.5 µg of anti TRPV1/isotype control, 45 min, 4C

↓ wash 2x (350 x g, 5min)

Stain with FITC-conjugated secondary antibody

↓

Incubate for 20 min, 4C

↓

wash 2x (350 x g, 5min), resuspend in PBS and run on Flow cytotmetry

↓

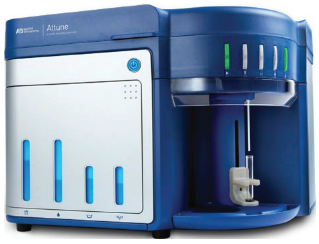
