## Supplementary material for "Optimized Flow Cytometric Detection of Transient Receptor Potential Vanilloid-1 (TRPV1) in Human Hematological Malignancies": Sup 3-Tables

**Supplementary 2**

**Table S1.** General characteristics of patients with hematological malignancies

| | | |
|---|---|---|
| Age | Years (range) | 31- 85 |
| Sex | Male | 28 |
| | Female | 21 |
| Subjects | *De novo* | 12 |
| | Under treatment | 37 |
| | Control | 21 |
| Diagnosis | B-NHL | 28 |
| | MM | 12 |
| | Acute Monocytic Leukemia | 2 |
| | MPD | 4 |
| | HCL | 1 |
| | ALL | 1 |
| | PTCL/NOS | 1 |

MM: multiple myeloma; B-NHL: B- Cell Non-Hodgkin's lymphoma; MPD: Myeloproliferative Disorder; HCL: Hairy-Cell Leukemia; ALL: Acute Lymphoblastic Leukemia; PTCL-NOS: Peripheral T-cell lymphoma/not otherwise specified.

**Table S2.** Comparison of mean TRPV1 MFI in THP-1, U266B1 and U937 cell lines and normal leukocytes

| Cell line | n | Mean MFI ($10^5$) (Cell Line) | Mean MFI ($10^5$) (Normal Leukocytes) | Ratio | Change |
|---|---|---|---|---|---|
| **THP-1** | 4 | 2.38 | 0.97 | 2.5 | Increased |
| **U266B1** | 1 | 1.81 | 0.76 | 2.38 | Increased |
| **U937** | 1 | 1.19 | 0.76 | 1.57 | Unchanged |

MFI: Median Fluorescence Intensity; n= number of experiments.
