## Supplementary material for "Optimized Flow Cytometric Detection of Transient Receptor Potential Vanilloid-1 (TRPV1) in Human Hematological Malignancies": Sup 3-Fig1B-raw data Western blot

### Original Fig 1B (upper section):

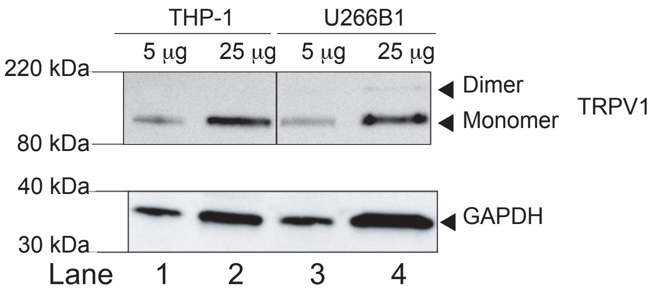


Fig 1B (upper section): raw WB data

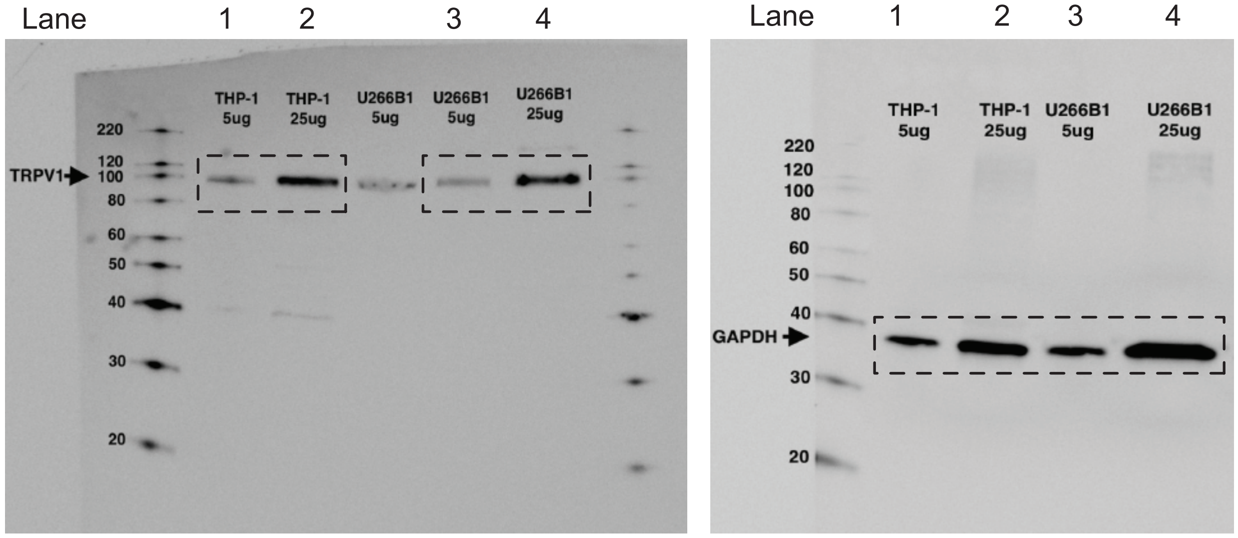


### Original Fig 1B (lower section):

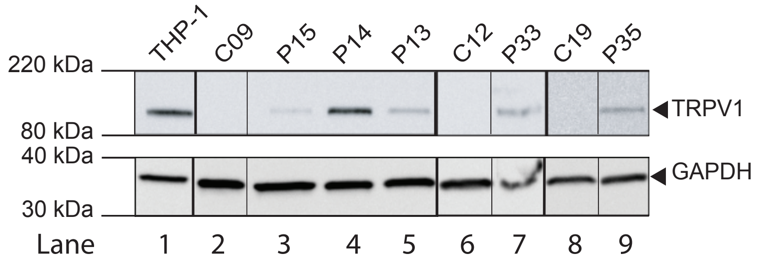


Fig 1B (lower section): raw WB data

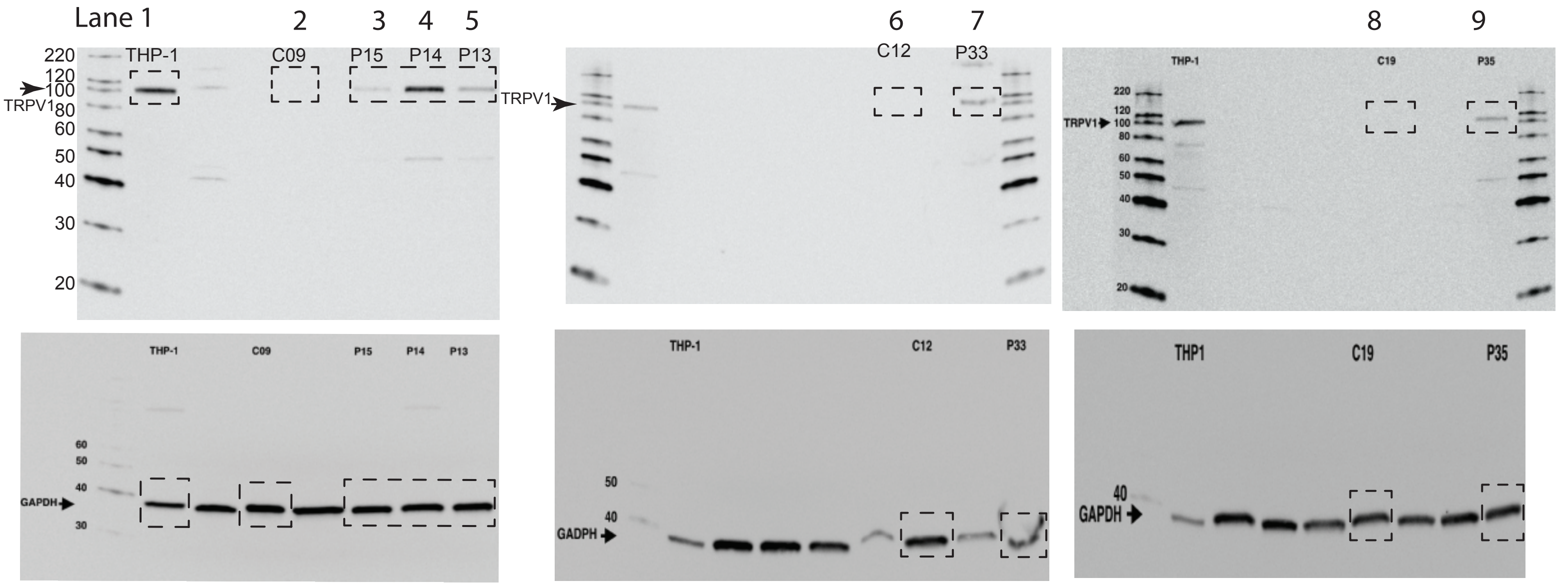
